# Long-Term Antibody Responses to *Vibrio cholerae* Across Natural Infection, Vaccination, and Challenge: A Systematic Review

**DOI:** 10.64898/2025.11.27.25341157

**Authors:** Aparna Mangadu, Christy Clutter, Kenadee Jacobson, Tatum Heiner, Shubhanshi Trivedi, Md. Saruar Bhuiyan, Ben J Brintz, Andrew S. Azman, Benjamin F. Arnold, Daniel T. Leung

## Abstract

**Background:** Cholera is a critical global health threat, with significant impact on low- and middle-income countries. Clinical surveillance is limited by non-specific symptoms, dependence on care-seeking, and availability of microbiological diagnostics, making burden estimation difficult. Serosurveillance is a potential solution, but the design and interpretation of cross-sectional seroepidemiological studies are currently limited by a lack of knowledge of longitudinal antibody trajectories.

**Methods:** We systematically reviewed literature from 1980 to 2025 containing longitudinal antibody measurements in humans following exposure to *Vibrio cholerae O1/O139* and associated vaccines.

**Results:** 51 studies representing adults and children from 1 low-income (1 study), 5 lower-middle-income (24 studies), 3 upper-middle-income (5 studies) and 5 high-income (21 studies) countries were included. 16 studies followed natural infection, 36 involved vaccination, and 6 were human challenge models, with some overlap. Antibody responses following infection were significantly higher than those induced by vaccination, particularly against O-specific polysaccharide (OSP). Younger children exhibited higher peak fold-change IgA responses to Cholera-toxin B-subunit (CTB) than older children and adults across infection and vaccine studies. Following vaccination, individuals from high-income countries showed higher IgA, IgG, and vibriocidal fold-change responses compared to those from lower-income settings. Limited data from challenge studies revealed slower decay of IgG compared to IgA and IgM.

**Conclusions:** This review enhances our understanding of longitudinal immune responses to *V. cholerae*, highlighting variability in antibody kinetics across age groups, geographic regions, and study designs, and informs the design and interpretation of seroepidemiological studies of cholera.

**Key Messages:**

- ur Research Question: We aimed to systematically review longitudinal antibody responses in humans following natural infection, vaccination, or challenge with *Vibrio cholerae* O1/O139, to inform interpretation of cross-sectional seroepidemiologic studies.
- We Found: We found that antibody responses vary significantly by exposure type, age group, and geographic setting, with infection inducing stronger and longer-lasting responses than vaccination, and younger children showing higher IgA responses to CTB compared to older children and adults.
- it is Important: These findings highlight variability in immune responses across populations and underscore the need for tailored approaches to accurately estimate cholera burden and guide global control strategies.

## Introduction

Seroepidemiology investigations use immunological markers from blood, primarily antibodies, which can be measured from population-based sampling and used to estimate population-wide infection burden and immunity following vaccination or natural infection^1^. Serosurveys could be an important tool for monitoring disease prevalence and transmission, identifying gaps in vaccine distribution or adherence, and strategizing public health interventions^2,3^. Despite its growing importance, a critical aspect of seroepidemiology is the need to examine the long-term kinetics of antibody responses to inform interpretations of single time-point measurements from individuals and populations in cross-sectional serosurveys.

Cholera remains a significant public health challenge, responsible for up to 4 million cases and 150,000 deaths annually^4,5^. Cholera particularly impacts those living in low- and middle-income countries due to inadequate health infrastructure, contaminated food and water sources, and insufficient disease surveillance and monitoring systems^6^. Clinical manifestations of *V. cholerae* infection vary from asymptomatic to severe acute watery diarrhea and vomiting, which can lead to severe dehydration and death due to hypovolemic shock^5^. The number of medically-attended cases likely represents only a small proportion of total infections, and serosurveillance, while not measuring clinical disease directly, offers a potential avenue for more accurate burden estimates^7^. However, interpretation of cross-sectional serosurveillance studies relies on modeling of antibody decay kinetics, which may differ depending on the population’s geographic location, age structure, and socioeconomic conditions. Additionally, specific to cholera seroepidemiology, a key question is whether antibody kinetics from one region, such as hyperendemic countries in Asia, can be generalizable to other regions, such as non-endemic countries, or endemic countries with infrequent outbreaks. Compiling data on long-term antibody persistence, or lack thereof, following *V. cholerae* exposure, across exposures, age groups, and socioeconomic conditions, can inform public health efforts against cholera worldwide.

This systematic review aims to collate and synthesize existing literature on global long-term antibody kinetics in humans following exposure to *V. cholerae*, with the goal of informing future seroepidemiological investigations and public health interventions.

## Methodology

### Overview

We conducted a systematic review to analyze existing literature that report on longitudinal antibody responses in humans after exposure to *Vibrio cholerae* through natural infection, vaccination, or controlled human challenge studies. Our methodology for this review was guided by the established systematic review framework developed by Khan and colleagues^10^, providing a structured approach for mapping key concepts and gaps in literature.

We conducted this review in four key stages: 1) identifying relevant studies through a comprehensive search of databases, 2) screening and selecting studies for full-text review based on specific inclusion and exclusion criteria (**Table 1**), 3) extracting relevant data from the included studies, and 4) collating, summarizing, and reporting the findings to highlight trends, patterns, knowledge gaps, and areas for future investigation related to long-term antibody kinetics.

**Table 1.** Inclusion and Exclusion Criteria.

| Study Characteristics | Inclusion Criteria | Exclusion Criteria |
| --- | --- | --- |
| **Study Design** | Randomized and non-randomized<br>Case only or case-control<br>Acute and convalescent<br>Pre-vaccine/post-vaccine<br>Pre-challenge/post-challenge<br>Point source outbreak with longitudinal follow up | Descriptive report or case-report without longitudinal follow-up<br>Prevalence, diagnostic, prognostic, or comparison study<br>Trial protocol<br>Only *in vitro* measurement |
| **Publication** | Peer reviewed journal<br>Published in English<br>Abstract and full-text available | Any academic dissertation<br>Conference proceedings, abstract, or poster<br>Any pre-print |
| **Pathogen** | *Vibrio cholerae* O1/O139 | Any other pathogen |
| **Specimen** | Serum or Plasma | Any other specimen |
| **Participants** | Humans of all ages | Non-human studies |
| **Interventions** | Natural Infection<br>Vaccine Trial<br>Challenge Model | Any other type of exposure or unable to determine exposure event |
| **Outcome Measures** | Longitudinal antibody and/or vibriocidal titers (follow-up $\geq 28$ days post onset of symptoms, final vaccine dose, or challenge event) | Antibody and vibriocidal titers not reported and/or no longitudinal follow-up |

### Search Strategy

We conducted a systematic literature search using the PubMed and Embase databases to identify relevant articles. Our search strategy included phrases created using specific key words: 1. Cholera, 2. Vibrio cholerae, 3. Antibody, and 4. Human. These terms were combined using Boolean operators to conduct search phrases such as ‘[“2” OR 1] AND 3’ and ‘[“2” OR 1] AND 3 AND MeSH Terms [4]’, where numbers correspond to the key terms listed above. We included Medical Subject Headings (MeSH) terms to ensure inclusion of articles indexed under standardized subject headings. We used this strategy to identify articles from the earliest available time up to and including April 2025. A total of 4,346 articles were identified and exported to Covidence online software where duplicates were removed.

### Selection Criteria, Data Management, and Extraction

We used the Covidence online platform for screening, full text review, and data extraction. After duplicates were removed, 2,179 articles were available for title and abstract screening. We reviewed articles based on specific inclusion and exclusion criteria (**Table 1**).

Two independent reviewers (MSB & ST or ST & AM) completed title and abstract screening using the inclusion criteria to ensure relevant studies were included for full-text review. We resolved any differences or discrepancies in inclusion/exclusion decisions between the two reviewers with discussion and consensus. Next, one independent reviewer (CC or AM) completed full-text reviews of included studies using the same inclusion and exclusion criteria (**Table 1**). Included studies from full-text reviews were then moved into the data extraction step. To ensure extraction of relevant data and to streamline the process, a data extraction template (**Supplementary Table 1**) was created by AM within the Covidence platform. The extraction template was used by two independent reviewers (AM & KJ or AM & TH) to gather and consolidate relevant data including: study design, country, country income status and intervention type. Income status was determined based on the most recent classifications by the World Bank. The extracted population-specific data included: inclusion and exclusion criteria, group differences, number of subjects, and population age. Finally, for each outcome measure, we extracted antibody titers at baseline, peak, and end of study, and seroconversion information as available. The independent reviewers who performed data extraction made antibody titer estimations and calculations based on information presented in the main text and figure captions. To reduce subjective bias, these extractions were compared and refined based on values derived with the online AI application, PlotDigitizer.

### Analysis

Extracted data were exported as a csv file and imported into R (Version 4.4.1) for analysis. Population sample sizes corresponding to each extracted value were used to calculate weighted means and associated confidence intervals. Because of the variability in measurement scales and units across studies, analyses focused on the fold change in the titer relative to baseline values. Fold change was defined as the ratio of the titer at each timepoint to the baseline titer. As these data were heavily right-skewed, weighted means and confidence intervals were estimated using nonparametric bootstrapping with 5,000 iterations. Using this bootstrapping approach, we estimated weighted mean peak titer fold changes for each antigen (e.g., OSP, LPS, CTB), stratified by isotype and intervention type. Additionally, we applied the same method to assess differences in peak titer fold changes across population age groups and country income levels. Weighted means of peak titer fold changes between intervention type, antigen, population age group, and country income type were compared using a one-way weighted ANOVA for variable with 3 or more independent groups and a two-sample t-test for variables with 2 independent groups using R, \**p*<0.05.

## Results

### Study Identification

We reviewed 2,179 abstracts published prior to and including April 2025 (**Figure 1**). Of those, 166 were selected for full-text review, and data from 51 studies were extracted and analyzed. Included studies were published between 1980 and 2024.

**Figure 1.**
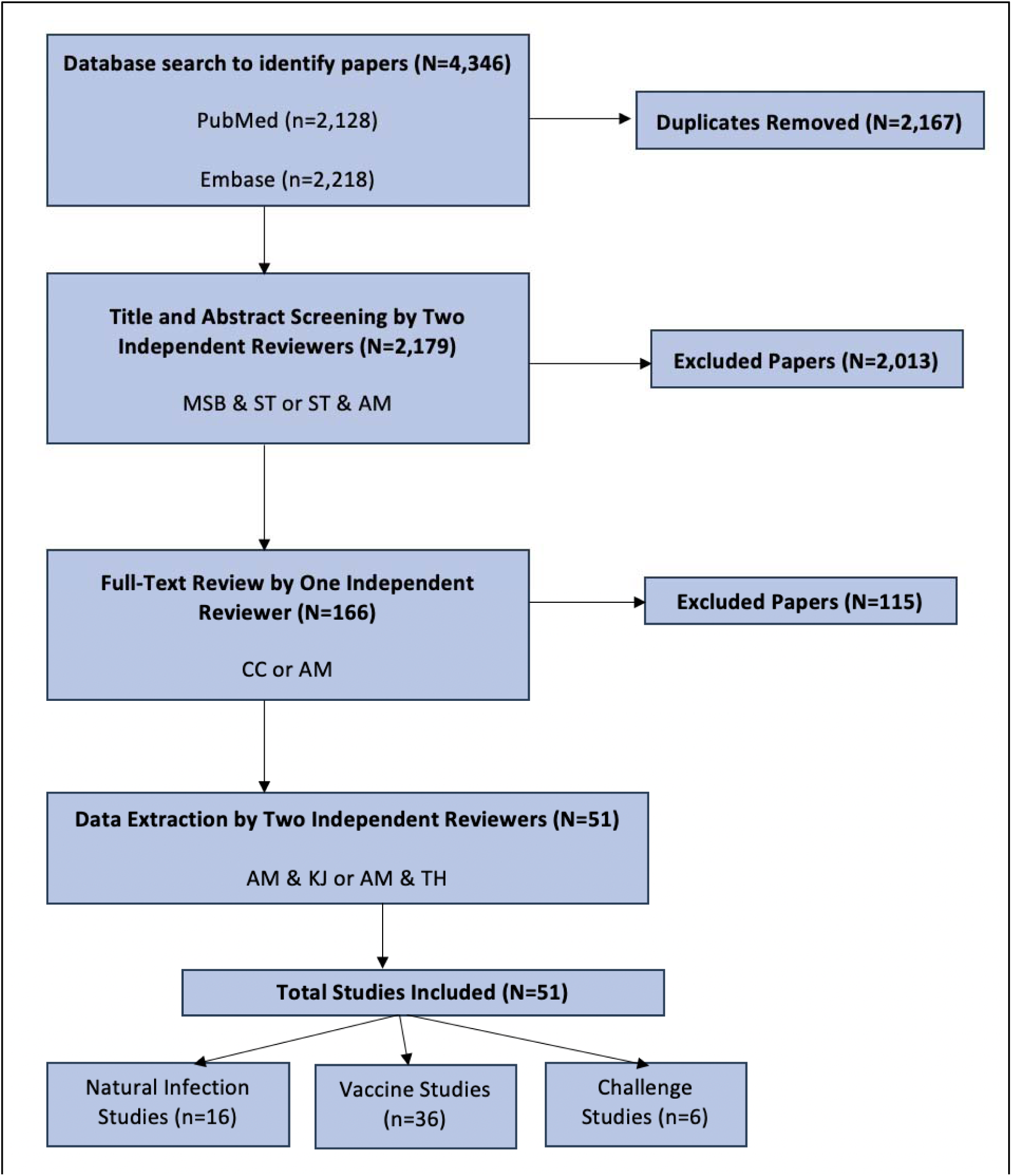
Flow diagram illustrating the selection of studies included in the systematic review A total of 4,346 records were identified through database searches (PubMed and Embase). After removal of 2,167 duplicates, 2,179 records underwent title and abstract screening by two independent reviewers, resulting in the exclusion of 2,013 records. Full-text review was conducted for 166 articles, of which 115 were excluded. Data extraction was completed for the remaining 51 studies by two independent reviewers. The final dataset included 51 studies: 16 natural infection studies, 36 vaccine studies, and 6 challenge studies.

**Figure 2.**
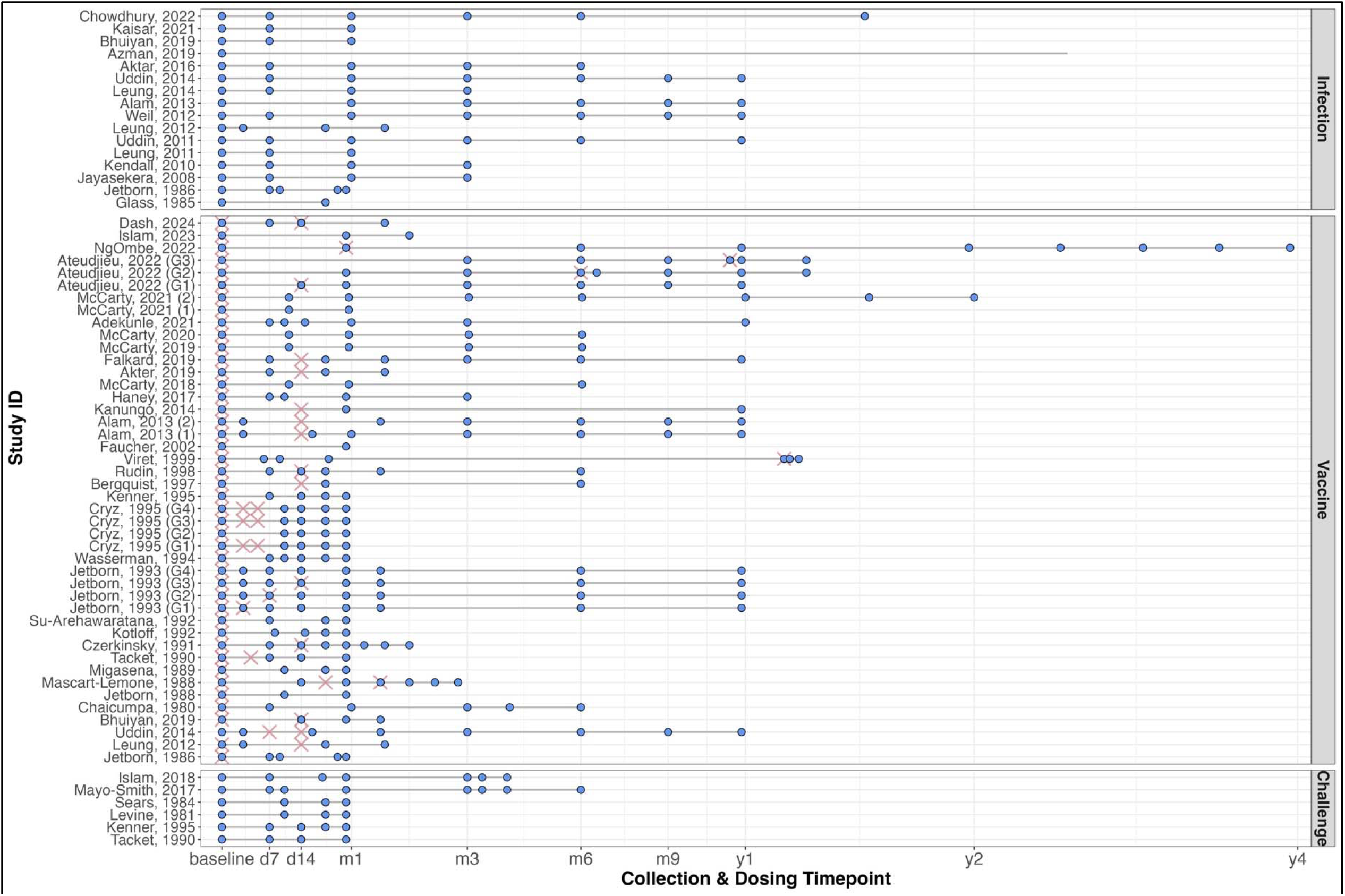
Sample collection timelines and vaccine dosing schedules across included studies Each horizontal line represents the timeline for a study (or study group), with blue circles indicating specimen collection timepoints and pink crosses indicating vaccine dosing events (vaccine studies only). Studies are grouped by study type (Challenge, Infection, Vaccine). Multiple rows for a single study denote different dosing or collection groups within that study. The x-axis (square root transformed) displays standardized timepoints from baseline through later follow-up (e.g., day 7, day 14, month 3, year 1, etc.). Grey horizontal lines provide a visual guide to the duration and structure of each study’s collection schedule.

The included were categorized into three types of pathogen exposure, 1) natural infection, 2) vaccination, and 3) human challenge model (**Supplementary Table 2**). Vaccine studies were further categorized into vaccines containing the Cholera Toxin B Subunit (CTB) and those without CTB. The primary assay used throughout included studies was an enzyme-linked immunosorbent assay (ELISA). Several studies also used vibriocidal assays. These assays were used to measure immunoglobulin antibody levels (i.e. IgG, IgA, and IgM) to various antigens (e.g. LPS, OSP) as well as vibriocidal responses in human serum and/or plasma.

Antigen type significantly influenced peak antibody responses across study types (**Table 2**). Natural infection studies showed significantly higher fold increases in IgG, IgA, and IgM titers to O-Specific Polysaccharide (OSP) compared to vaccine and challenge studies (IgG: *p* = 0.0019; IgA: *p*=0.0004; IgM: *p* = 0.0013).

**Table 2.** Weighted Peak Titer Comparisons Across Antigens by Isotype and Study Type.

| Antigen | N<sub>eff</sub><sup>a</sup> | Peak Titer - Fold Change from Baseline<br>[Weighted Mean (95% CI)] |
| --- | --- | --- |
| **IgG** |  |  |
| **Natural Infection** |  |  |
| O-Specific Polysaccharide (OSP) | 3.6 | 5.7 (3.4 – 8.3) |
| Lipopolysaccharide (LPS) | 3.6 | 2.8 (2.2 – 3.7) |
| Cholera Toxin B Subunit (CTB) | 3.3 | 4.0 (3.5 – 4.6) |
| Cholera Toxin (CT) | 1.0 | 12.5 |
| Sialidase | 1.0 | 3.6 |
| **Non-CTB Containing Vaccines** |  |  |
| O-Specific Polysaccharide (OSP) | 3.4 | 2.1 (1.2 – 2.7) |
| Lipopolysaccharide (LPS) | 1.9 | 1.2 (1.1 – 1.3) |
| **CTB-Containing Vaccines** |  |  |
| O-Specific Polysaccharide (OSP) | 5.6 | 1.6 (1.1 – 2.2) |
| Lipopolysaccharide (LPS) | 6.6 | 2.5 (2.0 – 2.9) |
| Cholera Toxin B Subunit (CTB) | 16.4 | 6.6 (4.6 – 8.6) |
| **Challenge** |  |  |
| O-Specific Polysaccharide (OSP) | 3.9 | 2.0 (1.2 – 3.2) |
| Cholera Toxin B Subunit (CTB) | 2.4 | 6.8 (6.2 – 8.3) |
| Cholera Toxin (CT) | 1.0 | 1.5 |
| Toxin co-regulated pilus A (TcPA) | 1.9 | 1.6 (2.5 – 1.8) |
| **IgA** |  |  |
| **Natural Infection** |  |  |
| O-Specific Polysaccharide (OSP) | 3.6 | 13.2 (6.2 – 18.6) |
| Lipopolysaccharide (LPS) | 9.9 | 12.8 (8.6 – 17.6) |
| Cholera Toxin B Subunit (CTB) | 3.6 | 10.6 (8.3 – 13.5) |
| Sialidase | 2.0 | 1.9 (1.1 – 2.7) |
| **Non-CTB Containing Vaccines** |  |  |
| O-Specific Polysaccharide (OSP) | 3.3 | 2.6 (2.0 – 2.9) |
| Lipopolysaccharide (LPS) | 1.9 | 1.3 (1.3 – 1.4) |
| **CTB-Containing Vaccines** |  |  |
| O-Specific Polysaccharide (OSP) | 5.6 | 2.9 (1.7 – 4.9) |
| Lipopolysaccharide (LPS) | 6.6 | 3.9 (3.8 – 4.2) |
| Cholera Toxin B Subunit (CTB) | 14.5 | 13.1 (9.2 – 17.5) |
| **Challenge** |  |  |
| O-Specific Polysaccharide (OSP) | 3.9 | 2.9 (1.6 – 4.7) |
| Toxin co-regulated pilus A (TcPA) | 1.9 | 1.3 (1.2 – 1.5) |
| **IgM** |  |  |
| **Natural Infection** |  |  |
| O-Specific Polysaccharide (OSP) | 3.6 | 6.4 (3.5 – 9.7) |
| Lipopolysaccharide (LPS) | 1.9 | 3.1 (2.1 – 5.3) |
| Cholera Toxin B Subunit (CTB) | 1.1 | 1.09 (1.0 – 1.13) |
| **Non-CTB Containing Vaccines** |  |  |
| O-Specific Polysaccharide (OSP) | 3.4 | 1.6 (1.1 – 2.1) |
| **CTB-Containing Vaccines** |  |  |
| O-Specific Polysaccharide (OSP) | 5.6 | 1.8 (1.2 – 2.4) |
| Lipopolysaccharide (LPS) | 1.6 | 1.9 (0.95 – 2.5) |
| Cholera Toxin B Subunit (CTB) | 2.6 | 1.2 (1.0 – 1.04) |
| **Challenge** |  |  |
| O-Specific Polysaccharide (OSP) | 3.9 | 2.3 (1.7 – 2.9) |
| **Vibriocidal** |  |  |
| **Natural Infection** | 5.9 | 96.8 (78.3 – 133.9) |
| **Non-CTB Containing Vaccines** | 12.0 | 6.8 (4.7 – 9.8) |
| **CTB-Containing Vaccines** | 3.4 | 139.8 (115.0 – 163.0) |
| **Challenge** | 1.9 | 8.9 (2.3 – 16.3) |

<sup>a</sup> Effective Sample Size –defined as $n_{eff} = (\sum w_i)^2 / \sum w_i^2$ . This metric adjusts the raw sample size to reflect information loss due to variation in sampling weights (population sample size) and represents the size of an equivalent simple-random sample

### Natural Infection Studies

Among studies involving natural cholera infection (*n*=16), antibody titers typically peaked between 7- and 30-days after symptom onset, with most reaching their maximum at day 7. All studies involving natural infection were conducted in Bangladesh.

Age related differences were observed in IgA responses with children aged 0-5 years exhibiting higher peak fold increases compared to older children (5-18 years) and adults (**Supplementary Table 3**). This difference was antigen dependent. Peak IgA responses to CTB were significantly greater in children aged 0-5 years than in older age groups (*p* = 0.0441; **Supplementary Table 3**).

Longevity of antibody responses appeared to be influenced by isotype, with IgG having more gradual declines than IgA following peak response. 6 months after natural infection, the latest time point available for studies including children, fold changes from baseline titers were similar between children and adults (**Figure 3b**).

**Figure 3.**
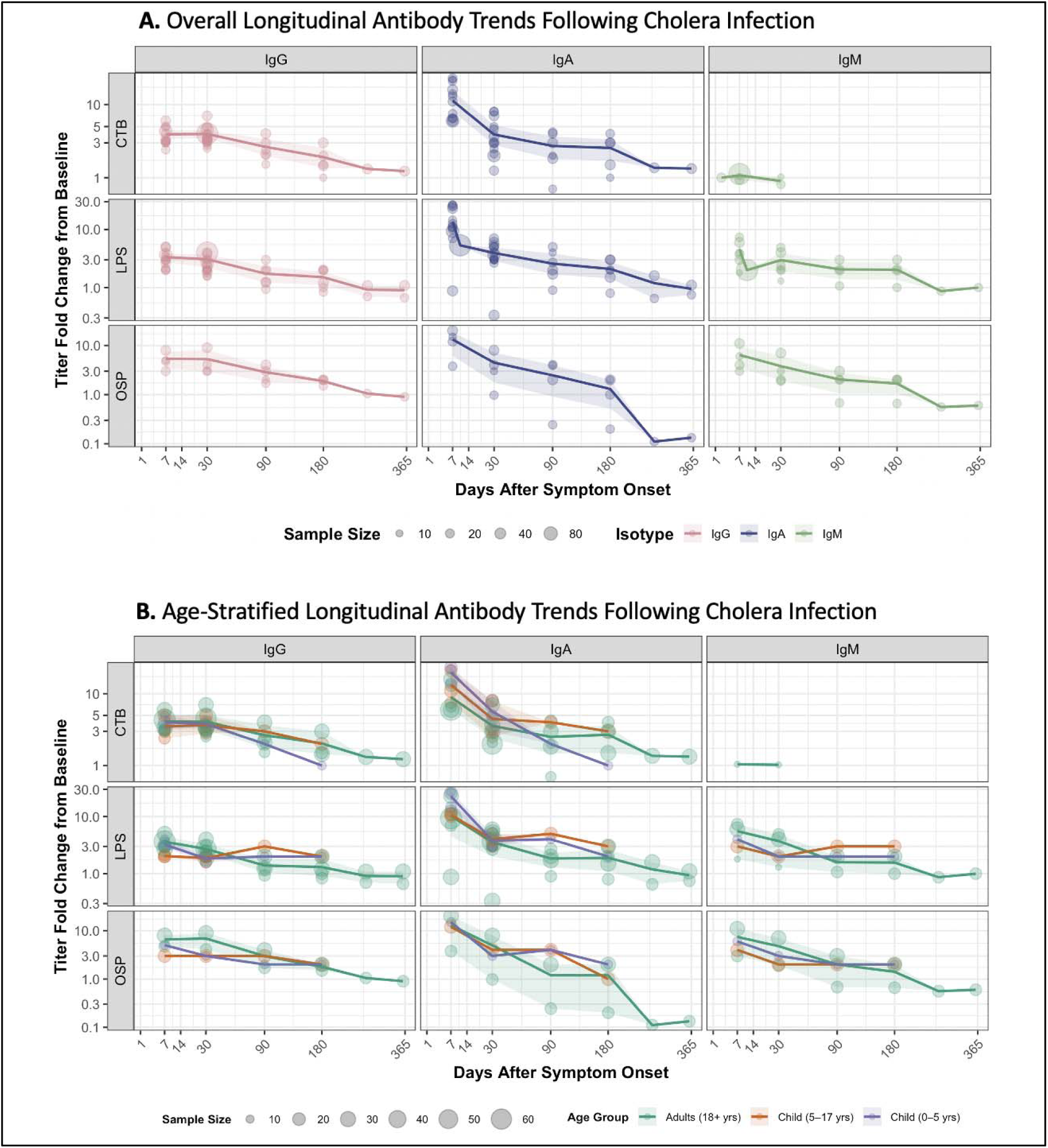
Longitudinal antibody responses in natural infection studies Longitudinal fold-change antibody responses from eligible natural infection studies are shown and stratified by isotype (IgA, IgG, IgM). **A.** Overall longitudinal trends. **B.** Age-stratified longitudinal trends. Each point represents an individual observation, with point size proportional to the underlying sample size. The dark solid line denotes the weighted mean response, and the shaded region indicates the 95% confidence interval across the 365 days following symptom onset.

### Vaccine Studies

A total of 36 vaccination studies were conducted across 14 countries, including Sweden (*n*=5), the United States (*n*=13), Bangladesh (*n*=9), Thailand (*n*=3), and others (Austria, Haiti, Colombia, India, Switzerland, Pakistan, Gabon, Australia, Cameroon, and Zambia; one study each). Vaccines evaluated included formulations containing CTB like Dukoral and Vaxchora, and vaccines without CTB like Shanchol, administered as single (*n*=19), two-dose (*n*=16), and three-dose (*n*=7) regimens.

Participants from high-income countries that received a vaccine containing CTB demonstrated substantially greater fold increases in peak vibriocidal antibody titers compared to those from upper-middle and lower-middle income countries (**Supplementary Table 4**). Similarly, IgG and IgA responses targeting the CTB antigen were markedly higher in high-income settings than lower-middle income countries among participants that received a vaccine containing CTB. Age also influenced immunogenicity with children aged 5-18 years exhibiting the highest peak fold increases in vibriocidal antibody responses compared to younger children and adults (*p* < 0.001; **Supplementary Table 3**). Longitudinal analysis revealed substantial variability in antibody kinetics across age groups and country income settings (**Supplementary Figure 1**).

### Human Challenge Studies

Limited data were available from studies involving exposure to *V. cholerae* through a human challenge model (*n*=6). Peak antibody titers were observed at 10-, 14-, and 28-days post challenge. All human challenge models were conducted in adult populations in the United States. Longitudinal trends indicated that IgG responses peak later than IgA and IgM responses (**Figure 4**).

**Figure 4.**
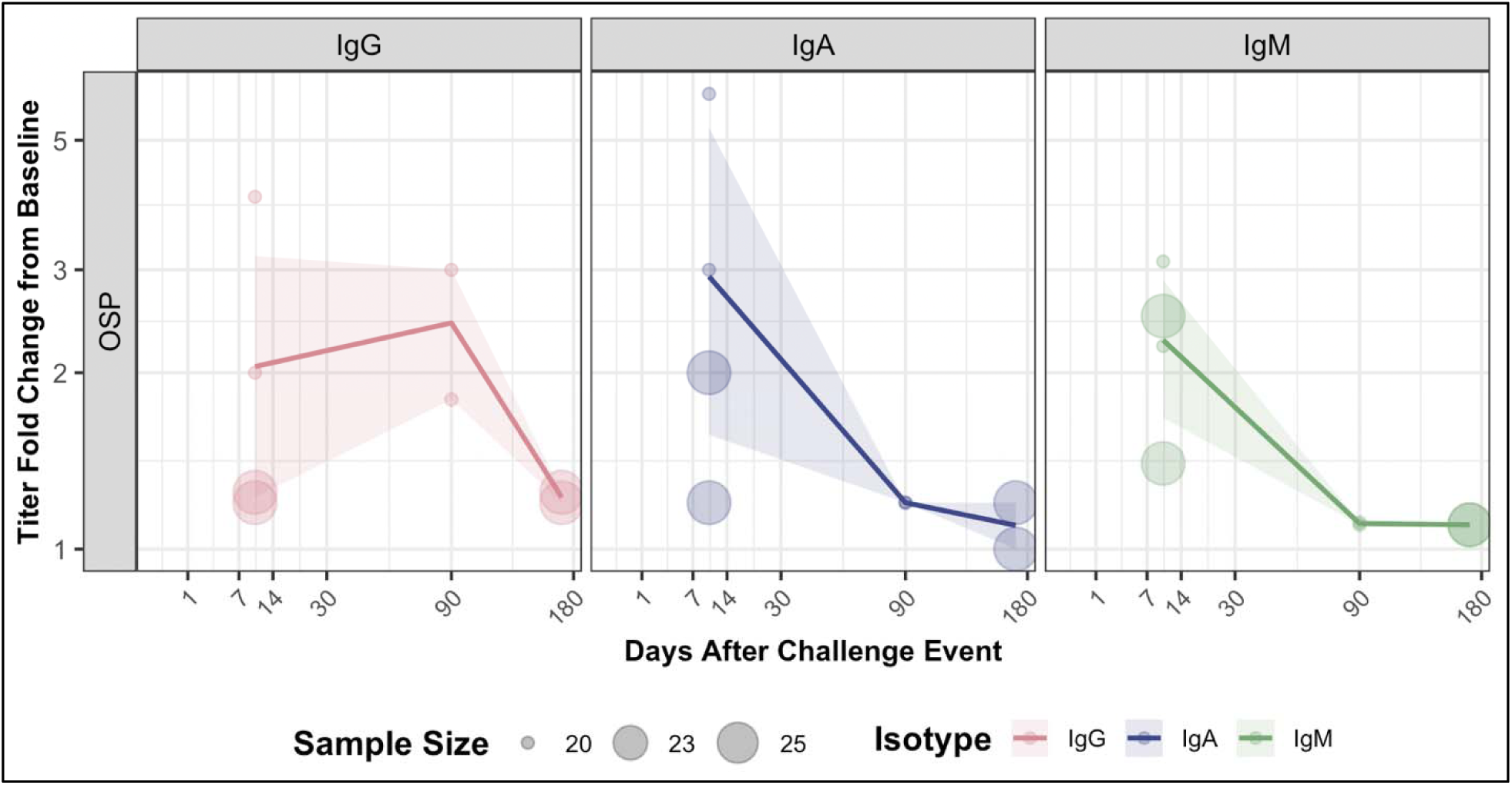
Longitudinal antibody responses in challenge studies Longitudinal fold-change antibody responses from challenge studies meeting inclusion criteria are shown and stratified by isotype (IgA, IgG, IgM). Each point represents an individual observation, with point size proportional to the contributing sample size; transparency is applied to enhance visualization of overlapping data. The dark solid line depicts the weighted mean response, and the shaded region represents the 95% confidence interval across the 182 days following the challenge event.

## Discussion

Serological surveillance is a promising, underutilized tool for estimating disease burden for bacterial pathogens like *Vibrio cholerae O1/O139*^7,11,12^. This systematic review synthesizes findings from 51 studies to characterize longitudinal antibody kinetics following natural cholera infection, vaccination, or experimental challenge. By examining how antibody responses vary across intervention types, population age, and country income level, this review provides insights that can improve the interpretation of cross-sectional serosurveys and inform public health strategies.

Our analysis shows notable age-related differences in immune responses, particularly for IgA antibodies. Children under 5 years of age exhibited significantly higher peak IgA responses to the CTB antigen following natural cholera infection, but not vaccination, compared to older children aged 5-18 years and adults, possibly due to lower baseline levels, and/or anamnestic response due to prior infection with heat-labile enterotoxin B subunit (LTB) of enterotoxigenic *Escherichia coli* (ETEC), which share structural and antigenic similarities with CTB^13^. A recent study similarly identified this trend, reporting substantial increases in IgA and IgG responses to CTB after infection but not vaccination^14^. Interestingly, older children demonstrated significantly higher fold changes in vibriocidal responses following vaccination compared to adults. Importantly, these age-related differences in antibody kinetics suggest that the age structure of a population must be considered when designing and interpreting serosurveys.

Antibody responses following natural cholera infection were consistently stronger than those elicited by vaccination, particularly for IgG, IgA, and IgM responses to the OSP antigen. This finding is consistent with prior studies^14^, underscoring the need to further investigate these differences to reduce seroincidence misclassification and inform other public health interventions. Further, the persistence of antibody responses varied by isotype in both natural infection and vaccine studies. As expected, IgG responses showed slower decay than IgA, presumably reflecting its longer half-life as a result of recycling through neonatal Fc receptor (FcRn) binding^15^. Vaccine-induced antibody peaks were more variable than those following natural infection, likely due to differences in vaccine formulation, dosing schedules (**Figure 2**), and administration routes. These factors complicate interpretation of seroepidemiologic findings and point to the need for standardized methodologies in vaccine trials.

Participants from high-income countries exhibited significantly higher peak fold-changes in IgG, IgA, and vibriocidal responses following administration of a vaccine containing CTB compared to those from lower-income settings. This disparity may reflect differences in baseline titer levels and health status, exposure history, gut microbiome composition, and nutritional factors^16–18^. These findings also emphasize the importance of considering contextual factors when interpreting vaccine performance across different populations.

The findings from this systematic review have several important implications for public health and future immunological investigations. First, there is a clear need for standardized protocols in measuring antibody responses^19^, including specific antigen targets and time points for sampling depending on use-case. Second, summarizing longitudinal antibody data can enhance our interpretation of cross-sectional serosurveys by providing critical context on seroprevalence and exposure history for specific populations. This data can help distinguish recent from past exposures and refine population-level disease burden and immunity estimates^19,20^. By characterizing antibody kinetics across age groups, interventions, and geographic settings, these results fill a methodological gap in seroepidemiology to support the development of more accurate and precise cholera burden estimation and transmission models.

While this review is comprehensive, there are several limitations to consider. First, several studies utilized figures for presenting antibody titers and concentrations rather than tables. We addressed this limitation by reducing subjective bias with an AI platform as described in the methods. Second, geographic representation was unevenly distributed, with natural infection studies exclusively from Bangladesh and challenge studies only from the United States. Further, challenge studies were only conducted in adult populations, limiting our ability to make comparisons across age groups. Finally, we are unable to make half-life estimations due to variability in antibody reporting between studies including differences in units, measures, and methods of measurement.

## Conclusion

Our work demonstrates that antibody responses to *V. cholerae* vary across populations, geographic regions, and interventions. By synthesizing global data on long term antibody kinetics in humans, this review provides a foundation for informing the design of future seroepidemiological studies.

## Data Availability

All data produced and used in the present study are available upon reasonable request to the authors.

## Ethics

This study used ONLY openly available human data derived from peer-reviewed publications identified through systematic searches of bibliographic databases (e.g., PubMed, Embase). All data were publicly available before the initiation of this work.

## Data Availability Statement

All data used in this systematic review were extracted from previously published studies. The compiled dataset generated during the review process is available from the corresponding author upon reasonable request.

## AI Tool Use Statement

Our use of AI was limited to the use of Plot Digitizer AI for reducing subjective bias in data extraction from figures.

