## Supplementary Table 1 for "Long-Term Antibody Responses to *Vibrio cholerae* Across Natural Infection, Vaccination, and Challenge: A Systematic Review"

**Supplementary Table 1.** Data Extraction Template

| **Item** | **Input** |
| --- | --- |
| IDENTIFICATION | |
| Study Details-Sponsorship Source | Open-Ended |
| Study Details-Country | Open-Ended |
| Study Details-Setting | Open-Ended |
| Authorship-Primary Author’s Name | Open-Ended |
| Authorship-Institution | Open-Ended |
| Authorship-Email | Open-Ended |
| Authorship-Address | Open-Ended |
| Additional Identification-Start Date | Open-Ended |
| Additional Identification-End Date | Open-Ended |
| Additional Identification-Journal | Open-Ended |
| Additional Identification-Year Published | Open-Ended |
| Additional Identification-Country Income Type | Select one:   - Low - Lower-middle - Upper-middle - High |
| METHODS | |
| Design | Select one:   - Randomized controlled trial - Case-control study - Cluster randomized controlled trial - Controlled interrupted time series - Historically controlled trial - Prospective cohort study - Retrospective cohort study - Other |
| Study Type | Select all that apply:   - Vaccine - Challenge - Infection |
| Specimen Type | Select all that apply:   - Serum - Plasma |
| Assay | Open-Ended |
| Isotype(s) | Select all that apply:   - IgG - IgA - IgM - Vibriocidal |
| Antigen(s) | Select all that apply:   - Sialidase - TcPA - CTB - OSP - LPS - CT |
| Seroconversion Citeria | Open-Ended |
| Days of Specimen Collection | Open-Ended |
| POPULATION | |
| Inclusion Criteria | Open-Ended |
| Exclusion Criteria | Open-Ended |
| Group Differences | Open-Ended |
| Number of Subjects | Open-Ended |
| Withdrawals | Open-Ended |
| Baseline Characteristics | **Characteristic** \| **Study Group X** \| **Overall**; **Mean Age (years (range))** \| \|; **Male** \| \|; **Female** \| \| |
| INTERVENTIONS | |
| Select Interventions | - Vaccine - Placebo - Infection - Challenge - Control |
| Extract Data (for each intervention) | \| **Intervention**; **Number of participants allocated** \|; **Dose** \|; **Frequency** \|; **Other** \| |
| OUTCOMES | |
| Select Outcomes | - IgG Antibody Titer - IgA Antibody Titer - IgM Antibody Titer - Vibriocidal Antibody Titer |
| Extract Data (for each outcome) | \| **Outcome**; **Antigen** \|; **Timepoints** \|; **Units** \|; **Direction** \|; **Notes** \|; **QA Outcome Group** \| |
| RESULTS DATA | |
| Extract Data (for each intervention-outcome) | Extract as available:   - Titer value - Timepoint - Fold change - Seroconversion % - Seroconversion ratio |

**Supplementary Table 2.** Characteristics of included studies (*n=*51)

| **Study ID** | **Country** | **Aim** | **Intervention and Study Design** | **Study Population** | **Specimen** | **Specimen Collection Schedule** | **Assay** | **Key Findings^a^** | **Ref** |
| --- | --- | --- | --- | --- | --- | --- | --- | --- | --- |
| Bergquist et al., 1997 | Sweden | Assess the safety and immunogenicity of CTB intranasal vaccine at various doses | Vaccine – volunteers divided into three dose groups – 10ug/dose, 100ug/dose, 1000ug/dose – and vaccinated twice intranasally with a 2-week interval – follow up for 1 year  Dosing Schedule – d0, d14 | Healthy Swedish adult volunteers (N=45) | Serum | d0, d21, m6 | ELISA | Moderate and high dose groups had significant IgG and IgA antibody increases to CTB antigen. IgA titers declined significantly by 6 months and IgG remained elevated. Peak IgA responses were seen on day 21 and peak IgG responses were seen at month 6. | (1) |
| Cryz et al., 1995 | Austria | Assess the safety and immunogenicity of a combined cholera and typhoid vaccine | Vaccine – volunteers randomly divided into four groups – 3 doses of Ty21a vaccine, 1 dose of CVD 103-HgR, 2 doses of Ty21A-CVD 103-HgR and monovalent Ty21a vaccine, and 2 doses of monovalent Ty21a and 1 dose of bivalent Ty21a-CVD 103-HgR vaccine – follow up for 4 weeks  Dosing Schedule -  Group 1 – d1, d3, d5  Group 2 – d1  Group 3 – d1, d3, d5  Group 4 – d1, d3, d5 | Healthy adult volunteers from the Medical University of Vienna (N=185) | Serum | d1, d10, d14, d21, d28 | ELISA, Vibriocidal | IgA antibodies to S typhi LPS were the highest in the Ty21a vaccine group. IgG antibodies to S typhi LPS were comparable between groups. Vibriocidal responses were highest when CVD 103-HgR was given alone or combined with Ty21a. Peak vibriocidal responses were observed on day 14 across all groups. | (2) |
| Jertborn et al., 1988 | Sweden, Bangladesh | Examine antibody responses from oral/subcutaneous immunization with cholera B subunit whole cell vaccine (B+WCV) or from natural cholera disease | Vaccine & Infection – volunteers divided into four groups – 1 oral dose of B+WCV (groups 1-2), 1 subcutaneous dose of B-WCV (group 3), natural cholera infection (group 4) – follow up for 4 weeks  Dosing Schedule - d0 | Healthy adult Swedish volunteers and Bangladeshi patients with natural cholera infection (4 groups, 5-7 per group) | Serum | d0, d7-10, d25-d28 | ELISA | Oral immunization resulted in significant increases in antitoxin IgA1 and IgG1-4 subclasses. Peak IgG subclass titers occurred on day 10 or day 28 and remained elevated for months. IgA1 antitoxin titers peaked on day 10 and decreased significantly after. | (3) |
| Glass et al., 1985 | Bangladesh | Examine antibody responses in cholera patients and family contacts | Infection – cholera patients and family contact with asymptomatic or symptomatic infections – follow up for 36 months | Cholera patients (N=10) and family contacts (N=1658) | Serum | d1, d21, m18-m36 | ELISA, Vibriocidal | Vibriocidal and IgG antitoxin responses peaked on day 21 in cholera patients but returned to baseline by month 36. 50% of family contacts that developed cholera had a significant vibriocidal response. | (4) |
| Kenner et al., 1995 | United States | Assess the safety, immunogenicity, and efficacy of Peru-15 a live oral cholera vaccine candidate | Vaccine & Challenge – volunteers randomly assigned to two groups – 1 dose of Peru-15, or 1 dose of placebo – challenged 1 month later – follow up for 4 weeks  Dosing Schedule – d0 | Healthy adult volunteers from Washington, DC area (N=11) | Serum | d0, d7, d14, d21, d28 | ELISA, Vibriocidal | Peak vibriocidal titers occurred on day 14. Challenge resulted in seroconversion among all volunteers | (5) |
| Azman et al., 2019 | Bangladesh | Use machine learning to understand immunological markers of V cholerae O1 infection | Infection – culture confirmed cholera cases – follow up for 915 days | Cholera patients at icddr,b (N=320) | Plasma | d2, follow up for 915 days | ELISA, Vibriodical | Vibriocidal Inaba and Ogawa, IgG, IgA, and IgM to LPS, and IgG and IgA to CTB had significant increases between day 2 and day 7 after onset of infection. Peak titers were observed at day 7 mostly. IgG to LPS and CTB peaked on day 30. Vibriocidal antibodies had biphasic decay. | (6) |
| Islam et al., 2018 | United States | Assess antibody responses after vaccination with live attenuated oral cholera vaccine CVD 103-HgR | Challenge – volunteers vaccinated with 1 dose of CVD 103-HgR and either challenged with wild type V cholerae O1 El Tor Inaba N16961 on day 10 or day 90 – followup for 90-170 days after challenge | Healthy adult North American volunteers (N=46) | Serum | 10 day challenge-d0,d7,d10,d20,d38,d180  90 day challenge-d0, d7, d20, d28, d90, d100, d118 | ELISA, Vibriocidal | IgM to V cholerae O1 Inaba OSP increased significantly on day 7 after vaccination and remained elevated for 28 days with a peak on day 10. IgA to V cholerae O1 Inaba OSP increased by day 10 but was back at baseline by day 28. Challenge at day 10 strain did not significantly increase IgM or IgA to OSP, but challenge at day 90 did result in significant IgG, IgM, and IgA to OSP increases. | (7) |
| Uddin et al., 2011 | Bangladesh | Evaluated antibody responses after natural cholera infection | Infection – patients with acute cholera infection – follow up for 360 days | Adult cholera patients at icddr,b (N= 18) | Plasma | d2, d7, d30, d90, d180, d360 | ELISA, Vibriocidal | Vibriocidal responses peaked on day 7, remained elevated until day 90, and were back at baseline by day 180. IgG and IgA to CTB responses peaked at day 7 to day 30. IgA remained elevated to day 90 and IgG to day 180. | (8) |
| Uddin et al., 2014 | Bangladesh | Evaluate antibody responses after vaccination with WC-rBS Dukoral vaccine of natural cholera infection from V cholerae O1 Ogawa | Vaccine & Infection – 2 doses of WC-rBS vaccine with a two-week interval or cholera patients – follow up for 360 days  Dosing Schedule – d0, d14 | Healthy adult vaccinees (N=33) and cholera patients at icddr,b (N=17) | Plasma | patients - d2, d7, d30, d90, d180, d270, d360  vaccinees-d0, d3, d17, d42, d90, d180, d270, d360 | ELISA, Vibriocidal | Significant increases in IgA, IgG, and IgM to OSP and LPS by day 3 after vaccination and peaks at day 17 after vaccination, remaining elevated for 1 month after second dose. Antibody responses to natural infection at same time points were higher with peaks at day 7. | (9) |
| Mayo-Smith et al., 2017 | United States | Evaluate antibody responses to a challenge strain V cholerae O1 El Tor Inaba N16961 in participants previously vaccinated with CVD 103-HgR Vaxchora | Challenge – participants had received either 1 dose of CVD 103-HgR or a placebo and were then challenged either 10 or 90 days later – follow up for 180 days post vaccination | challenged volunteers enrolled in the phase 3 clinical trial by PaxVax (N=84) | Serum | 10 day challenge-d7, d10, d20, d38, d180  90 day challenge-d7, d10, d28, d90, d100, d118, d180 | ELISA | Significant increases in IgG to CTB by day 7 after vaccination. Elevated CTB antibody levels at all days post challenge but higher fold changes in placebo groups. | (10) |
| Leung et al., 2012 | Bangladesh | Evaluate acute-phase immune responses to an oral cholera vaccine (OCV Dukoral WC-rBS) in children | Vaccine & Infection – participants were children divided into two age groups – young children 2-5 years and older children 6-17 years – administered 2 doses of vaccine 14 days apart or children with cholera infection – follow up for 30 days post vaccination/post presentation  Dosing Schedule – d0, d14 | Healthy young Bangladeshi children from an urban informal settlement area (N=40) and age matched children with cholera (N=38) | Plasma | d0, d3, d21, d44 | ELISA, Vibriocidal | Vaccine recipients and age matched infection participants had similar vibriocidal baseline levels. Vibiriocidal responses peaked at day 7 after the second vaccine dose and were comparable between age groups at all time points. Children with cholera infection had higher vibriocidal titers than vaccinees. IgG and IgA to CTB and IgA, IgG, and IgM to LPS responses were significantly higher by day 7 in all vaccinees with older children having higher IgG titers to CTB. | (11) |
| Leung et al., 2011 | Bangladesh | Evaluate immune responses from wild-type V cholerae O1 infection in children and adults | Infection – patients with cholera infection divided into 3 age groups – young children 3-5 years, older children 6-17 years, and adults 18-60 years – follow up for 30 days | Patients with V cholerae O1 serotype Ogawa infection at icddr,b (N=102) | Plasma | d2,d7,d30 | ELISA, Vibriocidal | IgG and IgA to CTB and LPS titers were significantly higher by days 7 and 30 in all age groups but young children had higher IgG to CTB baseline levels than adults. Adults had higher IgA to LPS responses at day 30 compared to young children. Most patients had significant vibriocidal responses on day 7 and children had higher increases than adults. | (12) |
| Aktar et al., 2016 | Bangladesh | Evaluate immune responses from V cholerae O1 infection in children and adults | Infection – patients with cholera infection divided into 3 age groups – young children 2-5 years, older children 6-17 years, and adults 18-55 years – follow up for 180 days | Patients with V cholerae O1 infection at icddr,b (N=60) | Plasma | d2, d7, d30, d90, d180 | ELISA, Vibriocidal | Vibriocidal responses peaked in all aged groups on day 7 and remained elevated for older children and adults until day 180. IgG, IgA, and IgM to OSP and LPS responses peaked on day 7 in all age groups. IgA and IgG to CTB in children peaked on day 7. IgG to CTB in adults peaked on day 30. | (13) |
| Weil et al., 2012 | Bangladesh | Evaluate immune responses from cholera infection | Infection – prospective follow-up of patients with cholera infection – follow up for 360 days | Patients with cholera infection from an urban high risk cholera area in Dhaka (N=54) | Serum | d2, d7, d30, d90, d180, d270, d360 | ELISA, Vibriocidal | 30% of participants had seroconversion of vibriocidal titers between day 90 and day 360. No significant increases in IgG and IgA to CTB and LPS were reported. | (14) |
| Falkard et al., 2019 | Haiti | Evaluate immune responses after vaccination with BivWC cholera vaccine in Haitian adults | Vaccine – participants divided into three cohorts – each received two doses of the vaccine 14 days apart -- follow up for 360 days  Dosing Schedule – d0, d14 | Healthy adults in the Saint Nicolas Hospital (N=73) | Plasma | d0, d7, d21, d44, d90, d180, d360 | ELISA, Vibriocidal | 80% of participants had seroconversion for vibriocidal titers after vaccination by day 44. Vibriocidal peaks were observed on day 21 after the second vaccine dose and remained elevated for 1 year. IgG to OSP significantly increased after vaccination and remained elevated for a year. IgM to OSP peaked on day 21 and IgA to OSP peaked on day 7 and remained elevated only until day 44. | (15) |
| Alam et al., 2013 (2) | Bangladesh | Evaluate antibody responses following natural cholera infection or oral cholera vaccination with Dukoral | Infection & Vaccine – adults cholera patients and healthy adults from an urban field site in Dhaka given 2 doses of Dukoral 2 weeks apart – follow up for 360 days  Dosing Schedule – d0, d14 | Adult cholera patients from icddr,b (N=30) and healthy Bangladeshi adult vaccinees (N=30) | Plasma | Vaccinees - d0, d3, d17, d30, d90, d180, d270, d360  patients - d2, d30, d90, d180, d270, d360 | ELISA | IgA and IgG to CTB and LPS responses were similar in vaccinees and patients. CTB responses are more long lived than LPS. Antibody responses peaked on day 30 in cholera patients. | (16) |
| Kendall et al., 2010 | Bangladesh | Evaluate immune responses from V cholera infection in Bangladesh | Infection – prospective follow up of cholera patients – follow up for 30 days | Adults cholera patients from icddr,b (N=32) | Plasma | d2, d7, d30, d90 | ELISA, Vibriocidal | Vibriocidal seroconversion occurred at day 7 in all subjects. IgG, IgM, and IgA to LPS were elevated at day 7 and day 30. | (17) |
| Jayasekera et al., 2008 | Bangladesh | Evaluate immune responses from V cholerae infection | Infection – prospective follow up of cholera patients -- follow up for 90 days | Adult cholera patients from icddr,b (N=15) | Serum | d2, d7, d30, d90 | ELISA, Vibriocidal | Vibriocidal seroconversion was observed in all patients by day 7 and remained elevated until end of follow up. 9 participants had peak IgA and IgG to CTB levels on day 7 and the other 5 had peak levels on day 30. IgG remained elevated and IgA returned to baseline. | (18) |
| Czerkinsky et al., 1991 | Sweden | Evaluate immune responses to vaccination with a newly developed oral cholera vaccine | Vaccine – participants divided to receive either two pectoral doses of the vaccine with a 2-week interval – follow up for 56 days from primary immunization  Dosing Schedule – d0, d14 | Healthy Swedish adult volunteers (N=30) | Serum | d0, d7, d14, d21, d28, d35, d44, d56 | ELISA | IgG and IgA to CTB peaked at day 21 post primary immunization and again at day 44 for individuals with 3 immunizations. | (19) |
| Akter et al., 2019 | Bangladesh | Evaluate immune responses to Shanchol oral cholera vaccine | Vaccine – participants administered two doses of Shanchol with a 2-week interval – follow up for 44 days (30 days post second immunization)  Dosing Schedule – d0, d14 | Healthy adult volunteers (N=30) | Plasma | d0, d7, d21, d44 | ELISA, Vibriocidal | Vibriocidal responses peaked at 7 days post the first vaccination and were not significantly impacted by a second dose. Seroconversion for vibriocidal responses occurred at all timepoints after day 7. IgG, IgA, and IgM to Ogawa OSP were significantly higher at day 7 and day 21 and decreased thereafter. Plasma antibody OSP specific responses were not boosted from a second dose of the vaccine. | (20) |
| Leung et al., 2014 | Bangladesh | Assess MAIT cell levels and B cell responses to natural cholera infection. Secondary – assess plasma antibody levels to natural cholera infection | Infection – prospective follow up of patients with V cholerae O1 infection – follow up for 90 days post onset of symptoms | Patients (adults and children) at icddr,b with stool culture positive for V cholerae O1 (N=23) | Plasma | d2, d7, d30, d90 | ELISA | IgG, IgA, and IgM in plasma to LPS peaked at day 7 and IgM and IgG were more long lived than IgA, IgA to CTB peaked at day 7 and declined thereafter. IgG to CTB continued to rise until day 30. IgM to CTB had gradual declines from baseline to day 30. | (21) |
| Levine et al., 1981 | United States | Evaluate immune responses to a rechallenge with cholera in volunteers that had previously had natural cholera infection | Challenge – cholera veterans and control subjects were challenged with V cholerae 33-36 weeks after prior infection – follow up for 28 days after challenge | Total participants (N=9) -- Cholera veterans (n=4) and controls subjects (n=5) | Serum | d0, d10, d21, d28 | ELISA, Vibriocidal | 2 out of 4 volunteers had significant increases in IgG antitoxic antibody following the challenge. | (22) |
| Wasserman et al., 1994 | Colombia, United States | Evaluate vibriocidal antibody responses to a live oral cholera vaccines CVD 103-HgR and CVD 110 using data from two sets of studies (preliminary studies and kinetics studies) | Vaccine – Preliminary studies were phase 1 and phase 2 clinical trials with North American volunteers given a single dose of CVD 103-HgR – follow up for up to 28 days. Kinetics studies included Colombian volunteers given a single dose of CVD 103-HgR, or North American volunteers given a single dose of CVD 103-HgR2 or CVD 110 – follow up for up to 28 days  Dosing Schedule - d0 | Total participants (N=307) -- Preliminary studies: North American volunteers (n=260) Kinetics studies: Colombian volunteers (n=27), North American volunteers (n=20) | Serum | d2, d7, d10, d14, d21, d28 | Vibriocidal | In preliminary studies, vibriocidal titers rose significantly between days 10 and 14. In kinetics studies, Colombian volunteers had peak vibriocidal responses on day 10, and day 10 also for North American volunteers. | (23) |
| Tacket et al., 1990 | United States | Assess safety, immunogenicity, and efficacy of hybrid typhoid-cholera vaccine strain EX645 against a V cholera O1 challenge | Vaccine & Challenge – participants received three doses of EX645 with 2-day intervals and were challenged with V cholerae one month later – follow up for 28 days after vaccination and challenge  Dosing schedule – d0, d2, d4 | Healthy adult volunteers vaccinated (N=14) and challenged (N=8) | Serum | d0, d7, d14, d28 after vaccine and challenge | ELISA, Vibriocidal | After vaccinations, 36% of volunteers had seroconversion by day 14. 1% of vaccinees had seroconversion for IgG to Inaba LPS and 7% for IgA to Inaba LPS. All challenged participants had seroconversion for vibriocidal responses. | (24) |
| Kanungo et al., 2014 | India | Assess the efficacy and immunogenicity of a modified killed whole cell oral cholera vaccine | Vaccine – participants randomized to receive 2 doses of the vaccine or placebo – follow up for 1 year  Dosing Schedule – d0, d14 | Healthy residents (adults and children) (N=137) | Serum | d0, d28, m12 | Vibriocidal | Vibriocidal titers to Inaba had significant increases 14 days after the first vaccine and declined thereafter. Vibriocidal rises were higher in younger children than adults at 14 days after the second dose. | (25) |
| Viret et al., 1999 | Switzerland | Evaluate immune responses after primary and booster immunization of V cholerae CVD 103-HgR | Vaccine – volunteers received a single dose of CVD 103-HgR and a booster dose 14 months later  Dosing schedule – d1, d418 | Healthy adults (N=5) | Serum | d0, d6, d9, d22, d418, d426, d439 | ELISA, Vibriocidal | Vibriocidal seroconversion occurred in 4/5 participants and 3/5 volunteers for IgG to CT. | (26) |
| Sears et al., 1984 | United States | Evaluate immune responses after a V cholerae challenge | Challenge – volunteers challenged with classical biotype of V cholerae Ogawa 395 or Inaba 569 or El Tor Inaba N16961 or El Tor Ogawa E7946 – follow up for 28 days after challenge | Healthy adult volunteers (N=114) | Serum | d0, d10, d21, d28 | ELISA | Participants challenged with strain Inaba had higher rises in antibody responses than strain Ogawa. | (27) |
| Migasena et al., 1989 | Thailand | Assess immunogenicity of vaccine candidate strain CVD 103-HgR | Vaccine – volunteers assigned to receive a single dose of the vaccine or placebo – follow up for 28 days  Dosing Schedule - d0 | Healthy Thai adult volunteers (N=24) | Serum | d0, d10, d21, d28 | ELISA, Vibriocidal | 92% of volunteers had significant rises in Inaba vibriocidal responses. | (28) |
| Mascart-Lemone et al., 1988 | Pakistan | Evaluate immune responses to combination of live oral typhoid vaccine and parenteral cholera vaccine | Vaccine – participants administered three doses of cholera and typhoid vaccines at 3-week intervals – follow up for 12 weeks  Dosing Schedule – d0, w3, w6 | Lactating mothers (N=6) | Serum | d0, w2, w4, w6, w8, w10, w12 | ELISA | Significant responses of IgA to V cholerae LPS was observed in 4/6 mothers. | (29) |
| Faucher et al., 2002 | Gabon | Assess immune response from combined CVD 103-HgR and Ty21a vaccine Gabonese children | Vaccine – participants received a single dose of vaccine – follow up for 28 days following vaccination  Dosing Schedule - d0 | Healthy school children from Lalala Public School receiving atovaquone/proguanil or placebo then vaccinated (N=330) | Serum | d0, d28 | Vibriocidal | Vibriocidal responses at baseline did not vary between vaccine and placebo and there was no statistical difference in vibriocidal response after vaccination. | (30) |
| Chaicumpa et al., 1980 | Thailand | Evaluate antibody responses after vaccination with a classical cholera vaccine | Vaccine – participants received either a single dose of the vaccine or sterile distilled water – follow up for 6 months after vaccination  Dosing Schedule - d1 | Male prisoners of Bangkok special prison hospital (N=20) | Serum | d1, d7, m1, m3, m4, m6 | Vibriocidal | Highest vibriocidal titers were observed day 7 after vaccination and remained elevated for 3 months. | (31) |
| McCarty et al., 2018 | Australia, United States | Assess the safety and immunogenicity of PXVX0200 CVD 103-HgR live attenuated recombinant V cholerae O1 vaccine | Vaccine – participants randomized to receive a single dose of the vaccine or placebo – follow up for 6 months after vaccination  Dosing Schedule - d1 | Healthy adult volunteers from 19 US and 6 Australian sites (N=3146) | Serum | d1, d11, d29, d181 | Vibriocidal | Serum vibriocidal seroconversion occurred by day 29 for vaccinees. Peak vibriocidal responses were observed on day 11 and decreased by day 181. | (32) |
| McCarty et al., 2019 | United States | Assess the safety and immunogenicity of PXVX0200 CVD 103-HgR live attenuated recombinant V cholerae O1 vaccine in older adults | Vaccine -- participants randomized to receive a single dose of the vaccine or placebo – follow up for 6 months after vaccination  Dosing Schedule - d1 | Adults aged 46-64 and other age groups from lot consistency study (N=389) | Serum | d1, d11, d29, d91, d181 | ELISA, Vibriocidal | Seroconversion in vibriocidal responses was higher in younger adults than older adults and seroconversion occurred by day 11 across all age groups. | (33) |
| Bhuiyan et al., 2019 | Bangladesh | Evaluate immune responses after natural cholera infection or oral cholera vaccine Shanchol | Vaccine & Infection – Participants either had natural cholera infection or two doses of heat killed cholera vaccine – follow up for 30 days (infection) and 42 days (vaccine)  Dosing Schedule – d0, d14 | Adult and pediatric patients with cholera infection (N=17) and health adult and pediatric vaccinees (N=17) | Plasma | Vaccine-d0, d14, d28, d42  infection - d2,d7, d30 | ELISA, Vibriocidal | Vibriocidal responses peaked at day 7 in cholera patients and day 14 in vaccinees. | (34) |
| Haney et al., 2017 | United States | Evaluate immune responses after vaccination with CVD 103-HgR vaccine | Vaccine -- Participants either received a single dose of the vaccine or placebo – follow up for 90 days  Dosing Schedule - d0 | Healthy adult participants (N=134) | Serum | d0, d7, d10, d28, d90 | Vibriocidal | Vibriocidal titers peaked on day 10 post vaccination. | (35) |
| Alam et al., 2013 (2) | Bangladesh | Compare immune responses to Dukoral killed whole cell oral cholera vaccine between adults and children | Vaccine – participants were stratified into age groups of young children (age 3.5-5 years), older children (age 7-14 years), and adults (age 20 to 45 years) and administered two doses of vaccine with a 2-week intervale – follow up for 42 days (28 days after second dose)  Dosing Schedule – d0, d14 | Healthy young children (N=20) older children (N=20), and adults (N=33) | Plasma | d0, d3, d42, d90, d180, d270, d360 | ELISA, Vibriocidal | Baseline IgA to CTB and LPS were comparable between adults and children but IgG to LPS and CTB baselines were higher in children than adults. Vibriocidal and IgG and IgA to CTB and LPS were significantly higher by day 3 after vaccination in all age groups but persisted for longer in adults than children. | (36) |
| Jertborn et al., 1993 | Sweden | Compare immune responses to B subunit whole cell cholera vaccine after two and three immunizations | Vaccine – volunteers were divided into four groups given 2 doses of vaccine at different time intervales, Group A 3 days, Group B 7 days, Group C 14 days, Group D 28-42 days – followed up for 12 months after the second dose  Dosing Schedule -  Group A: d0,d3  Group B: d0, d7  Group C: d0, d14  Group D: d0, d28-d42 | Healthy adult volunteers (N=65) | Serum | d3, d7, d14, d28, d42, m6, m12 | ELISA, Vibriocidal | 67% of Group A volunteers had significant IgA antitoxin titer increases but 100% for groups B-D. Vibriocidal responses did not increase in any group after the second dose. | (37) |
| Kotloff et al., 1992 | United States | Assess the safety and immunogenicity of CVD 103-HgR vaccine | Vaccine – Participants were randomized to receive a single dose of the vaccine or placebo – follow up for 28 days | Healthy college students (N=94) | Serum | d1, d8, d15, d21, d28 | ELISA, Vibriocidal | 97% of subjects experienced vibriocidal seroconversion after vaccination. | (38) |
| Jetborn et al., 1986 | Bangladesh | Evaluate immune responses to oral cholera vaccination or natural infection | Vaccine & Infection – samples analyzed from 4 trials with volunteers immunized with two or three doses of the vaccine or patients with natural cholera infection – follow up for 28 days after each injection or after onset of symptoms  Dosing Schedule - d1 | Healthy adult vaccinees (N=27) and cholera patients (N=20) | Serum | d0, d7, d9, d25, d28 after each dose | ELISA | Vaccinees had some increases in antitoxin antibody levels while clinical cholera patients had larger increases in both antitoxin and IgG to LPS antibody levels. Antitoxin antibody responses in both groups peaked between days 25-28. | (39) |
| McCarty et al., 2021 (1) | United States | Assess the safety and immunogenicity of PXVX0200 CVD 103-HgR vaccine | Vaccine –participants were divided into three cohorts and received a single dose of the vaccine – follow up for 29 days  Dosing Schedule - d1 | Children aged 2-5 years (N=399) and healthy adults (N=2688) | Serum | d1, d11, d29 | Vibriocidal | Across all cohorts, vibriocidal responses peaked on day 11. Responses were more persistent and long lived in adult participants. | (40) |
| Adekunle et al., 2021 | United States | Evaluate immune responses to CVD 103-HgR Vaxchora vaccine | Vaccine – participants administered a single dose of the vaccine – follow up for 365 days  Dosing Schedule - d1 | Healthy adult volunteers (N=12) | Serum | d1, d7, d10, d15, d30, d90, d365 | ELISA; Vibriocidal | Vibriocidal seroconversion occureed in all volunteers by day 10 and declined 46 fold by day 90. IgM and IgA to LPS peaked on days 10 and 15. | (41) |
| Chowdhury et al., 2022 | Bangladesh | Evaluate immune responses after natural cholera infection | Infection – cholera patients positive for V cholerae O1 – follow up for 540 days | Adult (N=25) and pediatric (N=25) cholera patients at icddr,b | Plasma | d2,d7,d30,d90,d180,d540 | ELISA, Vibriocidal | Anti-sialidase IgG titers peaked on day 30 while anti-sialidase IgA titers peaked on day 7 for adults. In children, titers increased from baseline to end of follow up. | (42) |
| Ateudijeu et al., 2022 | Cameroon | Evaluate immune responses to two doses of an oral cholera vaccine given at different intervals | Vaccine -- participants divided into 3 dose intervale groups (DIGs); DIG1 interval of 2 weeks, DIG2 interval of 6 months, and DIG3 interval of 11.5 months – follow up for 15 months  Dosing Schedule -  DIG 1: d1, d14  DIG 2: d1, m6  DIG 3: d1, m11.5 | Healthy volunteers (N=186) | Serum | d1,d14,d28,m3,m6,m6.5,m9,m11.5,m12,m15 | Vibriocidal | Vibriocidal responses were higher with a shorter interval period (DIG1) thank onger (DIG2 and DIG3). | (43) |
| NgOmbe et al., 2022 | Zambia | Assess immunogenicity of a 2-dose regimen of Shanchol oral cholera vaccine | Vaccine – participants received two doses of the vaccine – follow up for 48 months  Dosing Schedule – d0, d28 | Healthy adult volunteers (N=223) | Serum | d0, d28, m6, m12, m24, m30, m36, m42, m48 | Vibriocidal | Vibriocidal seroconversion was similar for both Inaba (25%) and Ogawa (26%) serotypes. Participants older than 34 years of age had higher odds of seroconversion. | (44) |
| Islam et al., 2023 | Bangladesh | Assess the safety and immunogenicity of a new oral cholera vaccine (OCV) and co administration with oral poliovirus vaccine (bOPV) | Vaccine – participants were randomized to receive either bOPV only, OCV only, or bOPV and OCV together – follow up for 56 days  Dosing Schedule - d0 | Healthy pediatric volunteers (N=579) | Serum | d0, d28, d56 | Vibriocidal | Participants that received the vaccine together had comparable vibriocidal titers to the groups that received bOPV or OCV alone. | (45) |
| Kaisar et al., 2021 | Bangladesh | Evaluate immune responses to natural cholera infection | Infection -- participants were either younger children (<5 years of age), older children (6-17 years of age), or adults (18-55 years of age) with V cholerae O1 El Tor biotype of Ogawa or Inaba infection – follow up for 30 days | Younger children (N=18), older children (N=29), and adults (N=31) with cholera | Plasma | d2, d7, d30 | Vibriocidal | Peak vibriocidal titers for both Ogawa and Inaba serotypes were observed on day 7 across all three age groups. | (46) |
| McCarty et al., 2021 (2) | United States | Evaluate immune responses to PXVX0200 CVD 103-HgR vaccine | Vaccine – participants administered a single dose of the vaccine – follow up for 730 days  Dosing Schedule - d1 | Healthy adolescent volunteers aged 12-17 years (N= 73) | Serum | d1,d11,d29,d91,d181,d365,d547,d730 | Vibriocidal | Vibriocidal Reponses peaked on day 11 and steadily declined until day 730. | (47) |
| Su-Arehawaratana et al., 1992 | Thailand | Assess the safety and immunogenicity of CVD 103-HgR | Vaccine – participants, either Royal Thai Army soldiers or civilians, were randomly assigned to receive one dose of the vaccine or placebo – follow up for 28 days  Dosing Schedule - d0 | Healthy Royal Thai Army soldiers (N=279) and civilians (N=120) | Serum | d0, d7, d21, d28 | Vibriocidal | 20-50% seroconversion was observed in soldiers after vaccination, but civilians expressed a higher vibriocidal response. | (48) |
| McCarty et al., 2020 | United states | Assess the safety and immunogenicity of PXVX0200 CVD 103-HgR vaccine | Vaccine –participants were divided into two cohorts and received a single dose of the vaccine – follow up for 180 days  Dosing Schedule - d1 | Healthy children and adolescents aged 6-17 years (N=321) | Serum | d1,d11, d29, d91, d181 | Vibriocidal | 100% seroconversion was observed by day 29 in cohort 1. Vibriocidal titers in cohort 1 peaked on day 11 and decreased until end of follow up. | (49) |
| Rudin et al., 1998 | Sweden | Evaluate immune responses in serum to oral cholera vaccine Dukoral | Vaccine – participants were divided into three groups and received either an oral or nasal cholera vaccine, one or two doses with a 2-week interval – follow up for 6 months  Dosing schedule – d0, d14 | Health Swedish volunteers aged 19-36 years old (N=27) | Serum | d0, w1, w2, w3, w6, m6 | ELISA | In volunteers vaccinated twice orally or nasally, IgG and IgG titers had significant increases by 7 days after the second dose. Single nasal dose volunteers had significant rises in IgG and IgA titers by 14 days after vaccination. | (50) |
| Dash et al., 2024 | Bangladesh | Evaluate immune responses to oral cholera vaccine Dukoral across different age groups | Vaccine – participants were divided into groups based on age, older children aged 5 to 18 years, younger children aged 2 to 5 years, and adults aged older than 18 years – administered two doses of vaccine with a 2-week interval – follow up for 44 days  Dosing Schedule – d0, d14 | Healthy older children (N=25), younger children (N=24), and adults (N=50) | Plasma | d-14,d0,d2,d7,d14,d44 | ELISA, Vibriocidal | Significant increases and seroconversion in vibriocidal titers were observed on day 7 after the first vaccine dose across all age groups and remained elevated for 1 month even after the second vaccine does. IgA and IgG to OSP was significantly increased by day 7 in older children. IgM to OSP significanlty increased by day 7 in younger children. IgG to OSP remained elevated for one month in adults and older children but IgA to OSP returned to baseline. | (51) |
| ^a^Key findings presented in this table are summarized in accordance with the objective of this scoping review and may not represent all key findings for each paper | | | | | | | | | |

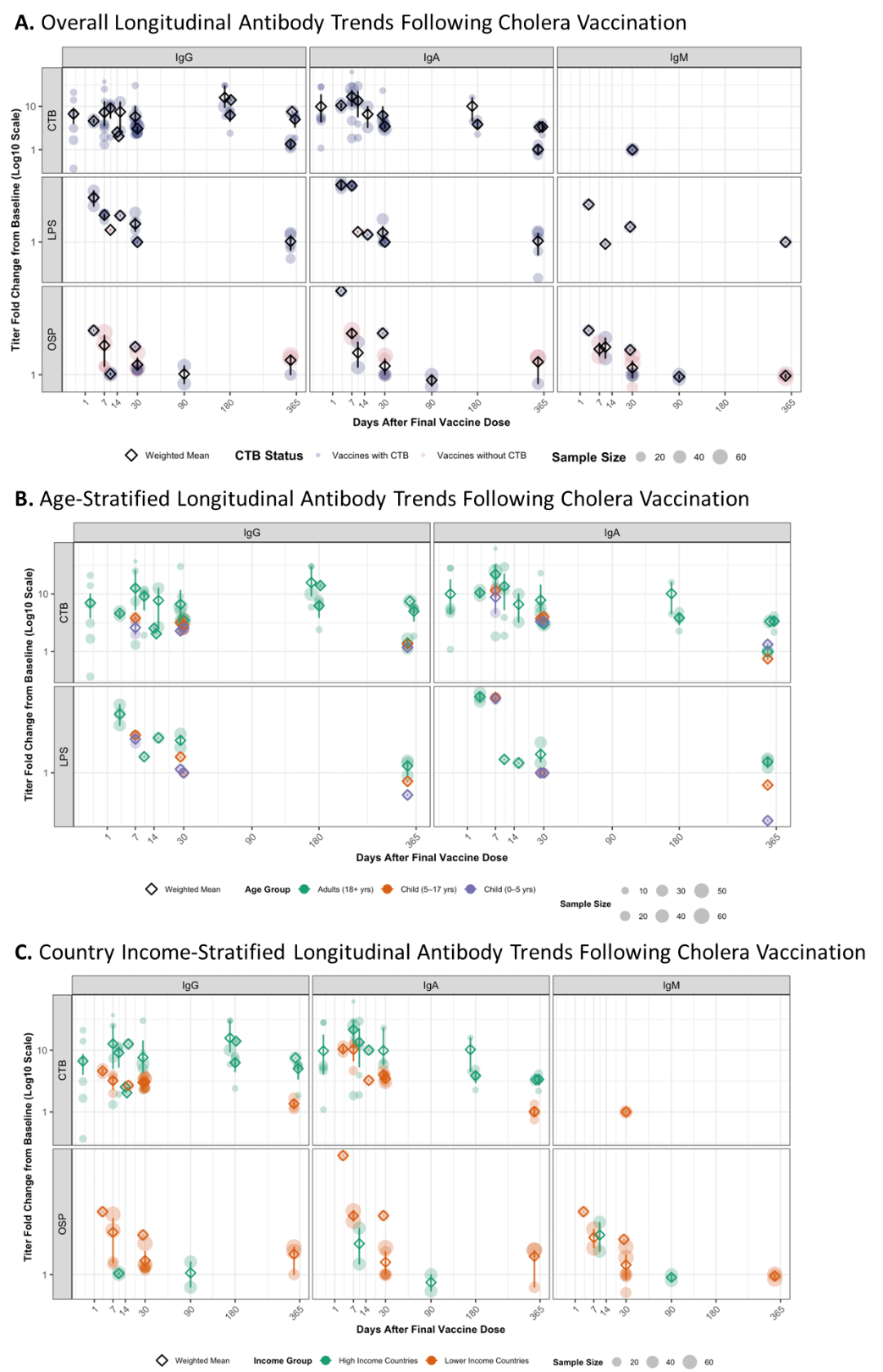

**Supplementary Figure 1.** Longitudinal antibody responses in vaccination studies

Longitudinal antibody responses were extracted from natural infection studies that met inclusion criteria. Responses are plotted as fold change from baseline titers and separated by antibody isotype (IgA, IgG, IgM). A. Overall longitudinal trends, B. Age-stratified longitudinal trends, C. Country Income-stratified longitudinal trends. Each point represents a raw data value, with point size proportional to sample size. Transparency was applied to individual points to visualize overlapping data. The empty diamond represents the weighted mean response per day, and error bars denote 95% confidence intervals over a 365-day period following symptom onset.

**Supplementary Table 3**. Weighted Peak Titer Comparisons Across Population Age by Isotype, Study Type, and Antigen

| **Antigen** | **Population age Group** | **N_eff_^a^** | **Peak Titer - Fold Change from Baseline**  **[Weighted Mean (95% CI)]** |
| --- | --- | --- | --- |
| IgG | | | |
| Natural Infection | | | |
| OSP | Younger Children (<5 Years) | 1.0 | 5.0 |
|  | Older Children (5-18 Years) | 1.0 | 3.0 |
|  | Adults (>18 Years) | 1.9 | 7.1 (4.8 - 9.0) |
| LPS | Younger Children (<5 Years) | 2.8 | 3.2 (2.9 - 4.0) |
|  | Older Children (5-18 Years) | 2.9 | 2.4 (2.0 - 3.0) |
|  | Adults (>18 Years) | 4.7 | 3.4 (2.7 - 4.1) |
| CTB | Younger Children (<5 Years) | 2.8 | 4.0 (3.2 - 5.0) |
|  | Older Children (5-18 Years) | 2.9 | 3.5 (2.4 - 5.0) |
|  | Adults (>18 Years) | 4.6 | 4.2 (3.2 - 5.4) |
| Sialidase | Adults (>18 Years) | 1.0 | 3.6 |
| Non-CTB Containing Vaccines | | | |
| OSP | Adults (>18 Years) | 3.4 | 2.1 (1.2 – 2.7) |
| LPS | Adults (>18 Years) | 1.9 | 1.2 (1.1 – 1.3) |
| CTB Containing Vaccines | | | |
| OSP | Younger Children (<5 Years) | 1.0 | 1.3 |
|  | Older Children (5-18 Years) | 1.0 | 1.7 |
|  | Adults (>18 Years) | 3.9 | 1.6 (1.0 – 2.4) |
| LPS | Younger Children (<5 Years) | 2.0 | 1.9 (1.7 – 2.0) |
|  | Older Children (5-18 Years) | 2.0 | 2.0 (2.0 – 2.0) |
|  | Adults (>18 Years) | 2.9 | 2.9 (2.4 – 3,5) |
| CTB | Younger Children (<5 Years) | 2.9 | 2.6 (2.0 – 3.2) |
|  | Older Children (5-18 Years) | 2.9 | 3.3 (2.4 – 4.0) |
|  | Adults (>18 Years) | 10.7 | 8.1 (5.7 – 10.9) |
| Challenge | | | |
| CT | Adults (>18 Years) | 1.0 | 1.5 |
| OSP | Adults (>18 Years) | 3.9 | 2.0 (1.2 - 3.2) |
| CTB | Adults (>18 Years) | 2.4 | 6.8 (6.2 - 8.3) |
| TcPA | Adults (>18 Years) | 1.9 | 1.6 (1.5 - 1.8) |
| IgA | | | |
| Natural Infection | | | |
| OSP | Younger Children (<5 Years) | 1.0 | 15.0 |
|  | Older Children (5-18 Years) | 1.0 | 12.0 |
|  | Adults (>18 Years) | 1.9 | 12.9 (3.8 - 20.0) |
| LPS | Younger Children (<5 Years) | 2.8 | 22.4 (14.5 - 27.0) |
|  | Older Children (5-18 Years) | 2.9 | 10.7 (10.0 - 11.0) |
|  | Adults (>18 Years) | 4.7 | 9.4 (4.5 - 15.3) |
| CTB | Younger Children (<5 Years) | 2.8 | 19.8 (14.0 - 23.0) |
|  | Older Children (5-18 Years) | 2.9 | 13.2 (7.5 - 21.0) |
|  | Adults (>18 Years) | 4.6 | 8.0 (5.3 - 12.1) |
| Sialidase | Adults (>18 Years) | 1.0 | 2.7 |
| Non-CTB Containing Vaccines | | | |
| OSP | Adults (>18 Years) | 3.4 | 2.6 (2.0 – 2.9) |
| LPS | Adults (>18 Years) | 1.9 | 1.3 (1.2 – 1.4) |
| CTB Containing Vaccines | | | |
| OSP | Younger Children (<5 Years) | 1.0 | 2.0 |
|  | Older Children (5-18 Years) | 1.0 | 2.0 |
|  | Adults (>18 Years) | 3.9 | 3.2 (1.5 – 6.2) |
| LPS | Younger Children (<5 Years) | 2.0 | 3.9 (3.8 – 4.0) |
|  | Older Children (5-18 Years) | 2.0 | 4.0 (4.0 – 4.0) |
|  | Adults (>18 Years) | 2.9 | 4.0 (3.7 – 4.4) |
| CTB | Younger Children (<5 Years) | 2.9 | 7.1 (4.0 – 13.0) |
|  | Older Children (5-18 Years) | 2.9 | 9.9 (7.0 – 12.5) |
|  | Adults (>18 Years) | 8.9 | 13.8 (8.6 – 19.9) |
| Challenge | | | |
| OSP | Adults (>18 Years) | 3.9 | 2.9 (1.6 - 4.8) |
| TcPA | Adults (>18 Years) | 2.9 | 1.3 (1.2 - 1.5) |
| IgM | | | |
| Natural Infection | | | |
| OSP | Younger Children (<5 Years) | 1.0 | 6.0 |
|  | Older Children (5-18 Years) | 1.0 | 4.0 |
|  | Adults (>18 Years) | 1.9 | 7.4 (3.0 - 11.0) |
| LPS | Younger Children (<5 Years) | 1.0 | 4.0 |
|  | Older Children (5-18 Years) | 1.0 | 3.0 |
|  | Adults (>18 Years) | 2.8 | 4.7 (1.8 - 7.3) |
| Non-CTB Containing Vaccines | | | |
| OSP | Adults (>18 Years) | 3.4 | 1.6 (1.1 – 2.1) |
| CTB Containing Vaccines | | | |
| OSP | Younger Children (<5 Years) | 1.0 | 1.2 |
|  | Older Children (5-18 Years) | 1.0 | 1.2 |
|  | Adults (>18 Years) | 3.9 | 1.9 (1.3 – 2.7) |
| LPS | Adults (>18 Years) | 1.6 | 1.9 (0.95 – 2.5) |
| CTB | Younger Children (<5 Years) | 1.0 | 1.0 |
|  | Older Children (5-18 Years) | 1.0 | 1.0 |
|  | Adults (>18 Years) | 1.0 | 1.04 |
| Challenge | | | |
| OSP | Adults (>18 Years) | 3.9 | 2.3 (1.7 - 2.9) |
| Vibriocidal | | | |
| Natural Infection | | | |
| - | Younger Children (<5 Years) | 4.8 | 208.2 (72.3 – 391.6) |
|  | Older Children (5-18 Years) | 4.8 | 142.6 (103.4 – 209.2) |
|  | Adults (>18 Years) | 5.6 | 92.0 (58.1 – 121.2) |
| Non-CTB Containing Vaccines | | | |
| - | Younger Children (<5 Years) | 4.0 | 5.0 (4.0 – 7.0) |
|  | Adults (>18 Years) | 4.7 | 7.8 (4.3 – 9.9) |
| CTB Containing Vaccines | | | |
| - | Younger Children (<5 Years) | 2.9 | 83.4 (7.5 – 158.7) |
|  | Older Children (5-18 Years) | 5.1 | 226.8 (141.0 – 260.5) |
|  | Adults (>18 Years) | 3.1 | 117.9 (55.4 – 135.3) |
| Challenge | | | |
| - | Adults (>18 Years) | 1.9 | 8.9 (2.3 – 16.3) |

**^a^** Effective Sample Size –defined as $n_{eff}={(\Sigma w_{i})}^{2}/\Sigma w_{i}^{2}$ . This metric adjusts the raw sample size to reflect information loss due to variation in sampling weights (population sample size) and represents the size of an equivalent simple‑random sample

**Supplementary Table 4**. Weighted Peak Titer Comparisons Across Country Income Type by Isotype, Study Type, and Antigen

| **Antigen** | **Country Income Type** | **N_eff_^a^** | **Peak Titer - Fold Change from Baseline**  **[Weighted Mean (95% CI)]** |
| --- | --- | --- | --- |
| IgG | | | |
| Natural Infection | | | |
| OSP | Lower Middle Income | 3.6 | 5.7 (3.4 - 8.3) |
| LPS | Lower Middle Income | 3.6 | 2.8 (2.2 - 3.7) |
| CTB | Lower Middle Income | 3.3 | 4.0 (3.5 - 4.6) |
| CT | Lower Middle Income | 1.0 | 12.5 |
| Sialidase | Lower Middle Income | 1.0 | 3.6 |
| Non-CTB Containing Vaccines | | | |
| OSP | Lower Middle Income | 3.4 | 2.1 (1.2 – 2.7) |
| LPS | High Income | 1.9 | 1.2 (1.1 – 1.3) |
| CTB Containing Vaccines | | | |
| OSP | High Income | 2.0 | 1.02 (1.0 – 1.04) |
|  | Lower Middle Income | 3.6 | 2.0 (1.5 – 2.7) |
| LPS | Lower Middle Income | 6.6 | 2.5 (1.9 – 2.9) |
| CTB | High Income | 7.9 | 10.8 (8.1 – 13.7) |
|  | Lower Middle Income | 8.0 | 3.5 (2.9 – 4.1) |
|  | Upper Middle | 1.0 | 1.9 |
| CT | High Income | 1.0 | 2.5 |
| Challenge | | | |
| OSP | High Income | 3.9 | 2.0 (1.2 - 3.2) |
| CTB | High Income | 2.4 | 6.8 (6.2 - 8.3) |
| CT | High Income | 1.0 | 1.5 |
| TcPA | High-Income | 1.9 | 1.6 (1.5 - 1.8) |
| IgA | | | |
| Natural Infection | | | |
| OSP | Lower-Middle Income | 3.6 | 13.2 (6.2 - 18.6) |
| LPS | Lower-Middle Income | 3.6 | 8.3 (3.1 - 16.3) |
| CTB | Lower Middle Income | 3.6 | 8.1 (5.2 - 13.5) |
| Sialidase | Lower Middle Income | 2.0 | 1.9 (1.1 - 2.7) |
| Non-CTB Containing Vaccines | | | |
| OSP | Lower Middle Income | 3.4 | 2.6 (2.0 – 2.9) |
| LPS | High Income | 1.9 | 1.3 (1.28 – 1.4) |
| CTB Containing Vaccines | | | |
| OSP | High Income | 2.0 | 1.7 (1.2 – 2.3) |
|  | Lower Middle Income | 3.6 | 3.7 (2.0 – 6.8) |
| LPS | Lower Middle Income | 6.6 | 3.9 (3.8 – 4.2) |
| CTB | High Income | 6.6 | 17.7 (11.1 – 24.0) |
|  | Lower Middle Income | 8.0 | 8.0 (5.5 – 10.6) |
| Challenge | | | |
| OSP | High Income | 3.9 | 2.9 (1.6 - 4.8) |
| TcPA | High Income | 1.9 | 1.3 (1.2 - 1.5) |
| IgM | | | |
| Natural Infection | | | |
| OSP | Lower Middle Income | 3.6 | 6.3 (3.5 - 9.7) |
| LPS | Lower Middle Income | 1.9 | 3.1 (2.1 - 5.3) |
| CTB | Lower Middle Income | 1.1 | 1.1 (1.10 - 1.1) |
| Non-CTB Containing Vaccines | | | |
| OSP | Lower Middle Income | 3.4 | 1.6 (1.1 – 2.1) |
| CTB Containing Vaccines | | | |
| OSP | High Income | 2.0 | 2.0 (1.5 – 2.5) |
|  | Lower Middle Income | 3.6 | 1.6 (1.1 – 2.6) |
| LPS | High Income | 1.0 | 0.95 |
|  | Lower Middle Income | 1.0 | 2.5 |
| CTB | Lower Middle Income | 2.6 | 1.02 (1.0 – 1.04) |
| Challenge | | | |
| OSP | High Income | 3.9 | 2.3 (1.7 - 2.9) |
| Vibriocidal | | | |
| Natural Infection | | | |
| - | Lower Middle Income | 5.9 | 97.1 (78.4 – 136.2) |
| Non-CTB Containing Vaccines | | | |
| - | High Income | 3.7 | 3.0 (2.0 – 4.1) |
|  | Lower Middle Income | 9.6 | 8.4 (6.1 – 11.9) |
|  | Low Income | 1.9 | 1.59 (1.55 – 1.6) |
| CTB Containing Vaccines | | | |
| - | High Income | 3.6 | 157.5 (139.0 – 207.6) |
|  | Upper Middle Income | 7.4 | 10.5 (3.1 – 25.7) |
|  | Lower Middle Income | 9.6 | 8.4 (7.2 – 9.9) |
| Challenge | | | |
| - | High Income | 1.9 | 8.9 (2.3 – 16.3) |

**^a^** Effective Sample Size –defined as $n_{eff}={(\Sigma w_{i})}^{2}/\Sigma w_{i}^{2}$ . This metric adjusts the raw sample size to reflect information loss due to variation in sampling weights (population sample size) and represents the size of an equivalent simple‑random sample
